# Evaluating a Widely Implemented Proprietary Deterioration Index Model Among Hospitalized COVID-19 Patients

**DOI:** 10.1101/2020.04.24.20079012

**Authors:** Karandeep Singh, Thomas S. Valley, Shengpu Tang, Benjamin Y. Li, Fahad Kamran, Michael W. Sjoding, Jenna Wiens, Erkin Otles, John P. Donnelly, Melissa Y. Wei, Jonathon P. McBride, Jie Cao, Carleen Penoza, John Z. Ayanian, Brahmajee K. Nallamothu

**Affiliations:** Dept. of Learning Health Sciences, University of Michigan Medical School, Ann Arbor, MI; Department of Internal Medicine, University of Michigan Medical School, Ann Arbor, MI; Division of Computer Science and Engineering, University of Michigan College of Engineering, Ann Arbor, MI; Department of Industrial and Operations Engineering, University of Michigan College of Engineering, Ann Arbor, MI; Dept. of Cellular and Molecular Biology, University of Michigan Medical School, Ann Arbor, MI; Department of Computational Medicine and Bioinformatics, University of Michigan Medical School, Ann Arbor, MI; Nursing Informatics, Michigan Medicine, Ann Arbor, MI; Institute for Healthcare Policy and Innovation, University of Michigan, Ann Arbor, MI

**Keywords:** coronavirus disease, deterioration index, predictive model, validation study

## Abstract

**Introduction:** The Epic Deterioration Index (EDI) is a proprietary prediction model implemented in over 100 U.S. hospitals that was widely used to support medical decision-making during the COVID-19 pandemic. The EDI has not been independently evaluated, and other proprietary models have been shown to be biased against vulnerable populations.

**Methods:** We studied adult patients admitted with COVID-19 to non-ICU care at a large academic medical center from March 9 through May 20, 2020. We used the EDI, calculated at 15-minute intervals, to predict a composite outcome of ICU-level care, mechanical ventilation, or in-hospital death. In a subset of patients hospitalized for at least 48 hours, we also evaluated the ability of the EDI to identify patients at low risk of experiencing this composite outcome during their remaining hospitalization.

**Results:** Among 392 COVID-19 hospitalizations meeting inclusion criteria, 103 (26%) met the composite outcome. Median age of the cohort was 64 (IQR 53-75) with 168 (43%) African Americans and 169 (43%) women. Area under the receiver-operating-characteristic curve (AUC) of the EDI was 0.79 (95% CI 0.74-0.84). EDI predictions did not differ by race or sex. When exploring clinically-relevant thresholds of the EDI, we found patients who met or exceeded an EDI of 68.8 made up 14% of the study cohort and had a 74% probability of experiencing the composite outcome during their hospitalization with a median lead time of 24 hours from when this threshold was first exceeded. Among the 286 patients hospitalized for at least 48 hours who had not experienced the composite outcome, 14 (13%) never exceeded an EDI of 37.9, with a negative predictive value of 90% and a sensitivity above this threshold of 91%.

**Conclusion:** We found the EDI identifies small subsets of high- and low-risk COVID-19 patients with fair discrimination. We did not find evidence of bias by race or sex. These findings highlight the importance of independent evaluation of proprietary models before widespread operational use among COVID-19 patients.

## INTRODUCTION

The coronavirus disease 2019 (COVID-19) pandemic is straining the capacity of hospitals and healthcare systems across the United States.^1,2^ Accurately identifying subgroups of COVID-19 patients at high- and low-risk for adverse outcomes could help to alleviate this strain by better directing scarce resources to those patients at greatest need. This need has led to the development and use of clinical prediction models in COVID-19 patients. Many of these models suffer from a high risk of bias due to sample sizes too small to allow for both model development *and* validation.^3^ Although the majority of studies have focused on newly developed models, many models already exist to detect clinical deterioration among hospitalized patients.

One of the most widely used models is the Epic Deterioration Index (EDI), which is implemented in hundreds of U.S. hospitals.^4^ The EDI was developed using data from 3 healthcare organizations between 2012 and 2016, and it uses clinical data to calculate risk scores at regular 15-minute intervals throughout a patient’s stay starting from the time of hospital admission. Although not specific to COVID-19 patients, the EDI has been widely used during the pandemic to support decision-making in COVID-19 patients.^5–7^

The widespread use of the EDI raises implementation concerns because there are no peer-reviewed publications describing its validity in any patient population. These concerns are particularly salient given that health systems are using the EDI in conflicting ways and with substantially different thresholds.^7^ Even prior to the onset of COVID-19, publicly available information about the EDI was limited to anecdotal reports of its value in critically ill patients.^8,9^ The proprietary nature of models such as the EDI make independent validation difficult due to a lack of complete information on the model’s functional form and parameters.^10^ However, independent evaluation is needed because hospital-based models often do not perform well in external validation studies and because the performance of models erodes over time as utilization patterns change.^11^ Additionally, some widely adopted proprietary models have previously been shown to be biased against African American patients even when race was not included as a predictor.^12,13^ Given that COVID-19 disproportionately impacts African Americans with respect to its incidence and complications, the validity of the EDI needs to be established generally and for vulnerable subpopulations.

These concerns have not prevented its use from being advocated.^5^ An Epic Systems spokesperson recently stated that “some hospitals are now using the model with confidence,”^6^ while others suggest it is “helping save lives.”^4^ In this context, we sought to independently validate the ability of the EDI to predict adverse outcomes among diverse patients hospitalized with COVID-19 at a large academic medical center. We also stratified our evaluation by race, sex, and age to evaluate the model performance among key subgroups of patients. Our findings have potential implications for how the EDI – currently deployed in hundreds of U.S. hospitals^4^ – may be validated and used by healthcare systems during the COVID-19 pandemic, and more broadly in how proprietary models should be evaluated.

## METHODS

### Study Cohort

Our study cohort included adults 18 years and older diagnosed with COVID-19 who were admitted to Michigan Medicine (i.e., the academic health system of the University of Michigan in Ann Arbor) between March 9, 2020 and May 20, 2020 from the emergency department, outpatient clinics, and outside hospital transfers. We excluded encounters where patients were admitted directly to an ICU (n=215), discharged to hospice (n=34), or where EDI scores were not available (n=10). We also excluded patients who remained hospitalized but had not yet experienced the composite outcome described below (n=27) because it was not possible to determine with certainty whether they would reach the primary outcome during their hospitalization. The study was approved by the Institutional Review Board of the University of Michigan Medical School.

### The Epic Deterioration Index Model

The EDI is generated from a proprietary early-warning prediction model developed by Epic Systems Corporation (Verona, WI) using data that are routinely recorded within its electronic health record. Epic is one of the largest healthcare software vendors in the world, and its electronic health record is used by most U.S. News and World Report’s top-ranked healthcare systems and reportedly includes medical records for nearly 180 million Americans (or 56% of the U.S. population).^14^

The EDI aims to detect patients who deteriorate and require higher levels of care. Its score ranges from 0 to 100, where higher numbers denote a greater risk of experiencing a composite adverse outcome of requiring rapid response, resuscitation, intensive care unit (ICU)-level care, or dying in the next 12-38 hours. Details related to the specific cohorts within which the model was developed, the model parameters, and its detailed performance characteristics have not been shared publicly or described in the published literature.

All hospitalized patients at Michigan Medicine have had calculations of the EDI as part of an ongoing evaluation of its clinical utility since late 2018; however, the EDI was not used in any clinical protocols during this time period. Calculations of the EDI begin immediately following hospital admission and then continue at regular 15-minute intervals until discharge. While the algorithm was developed prior to the COVID-19 pandemic, it includes several predictors that may be clinically relevant in COVID-19 patients: age, vital sign measurements (systolic blood pressure, temperature, pulse, respiratory rate, oxygen saturation), nursing assessments (Glasgow Coma Scale, neurological assessment, cardiac rhythm, oxygen requirement), and laboratory values (hematocrit, white blood cell count, potassium, sodium, blood pH, platelet count, blood urea nitrogen).

### Definition of the Primary Outcome

We defined our primary outcome as a composite of adverse outcomes that included the first of any of the following events that occurred during the hospitalization: ICU-level care, mechanical ventilation, or in-hospital death. We chose to include these adverse events for the composite outcome because they are highly relevant in the clinical care of COVID-19 patients where rapid respiratory decline is frequently described.

### Evaluation of the EDI to Identify High-Risk Patients

We used scores from the EDI calculated every 15 minutes throughout the hospitalization to predict the composite adverse outcome during the hospitalization. For patients who experienced the outcome, we only used EDI scores calculated prior to the outcome. We evaluated the discriminative performance of the EDI using the area under the receiver-operating-characteristic curve (AUC). The AUC represents the probability of correctly ranking two randomly chosen individuals (one who experienced the event and one who did not). Because the model runs every 15 minutes on all hospitalized patients, we calculated the AUC based on the entire trajectory of predictions using 2 strategies. First, we used the raw EDI scores to identify any time a patient crossed a decision threshold and deemed them ‘high-risk’ at that point, regardless of subsequent scores.^15^ This was done to reflect how such a model would likely be used in clinical practice to identify high-risk patients by flagging them for further clinical evaluation. For our second approach, we evaluated EDI slopes. Recent anecdotal reports have suggested that the rate of change of the EDI score may have predictive value that goes beyond the score itself.^16^ For these analyses, we evaluated the model discrimination using the EDI slope calculated on a rolling basis every 15 minutes using the prior 4 and 8 hours of EDI scores.

Predictions of deterioration are most beneficial when an appropriate lead time is available for action by clinicians. We therefore calculated a median lead time for the primary outcome by comparing when patients were first deemed high risk during their hospitalization to when they experienced the outcome. In all cases, we calculated empirical 95% confidence intervals (CI) for the AUC using 1,000 bootstrap replicates of our study cohort. Although it is unknown whether the EDI can be interpreted as a probability, model calibration was assessed using a calibration curve by comparing quintiles of all predicted EDI to the observed risk.

### Disparate Impact Analyses

To evaluate how the EDI performs in vulnerable populations, we conducted two analyses. First, we compared the AUC for defined by age (≥ 65 years versus < 65 years), sex, and race to determine if the EDI performs equally well in these subgroups. Then, we compared the the mean EDI in demographic subgroups and in those with and without the following comorbidities: cardiac arrhythmias, chronic kidney disease, chronic pulmonary disease, congestive heart failure, depression, diabetes mellitus, hypertension, liver disease, metastatic cancer, obesity, rheumatoid arthritis or other collagen vascular diseases, and solid tumors without metastases. This analysis was conducted to identify which comorbidities result in a higher EDI score, recognizing that comorbidities are not directly included in the EDI model.

### Evaluation of the EDI to Identify Low-Risk Patients 48 Hours after Admission

Another potential use of the EDI is to identify patients at low risk who could be sent home or to a lower-acuity setting, thereby offloading hospitals. Our goal was to evaluate how well the EDI at the end of 48 hours could identify patients at low risk of experiencing the outcome during the remainder of their hospitalization. We selected 48 hours following admission because decisions to triage patients to lower-acuity care within this timeframe may be valuable to hospital systems struggling in response to a surge of inpatient cases. For this analysis, we excluded patients who were discharged or experienced the composite outcome within the first 48 hours as triage decisions were not relevant for this group (n=65). We did this to remove very low-risk patients who were discharged as well as very high-risk patients who experienced the primary outcome early. The AUC was calculated based on the maximum EDI in the first 48 hours, with 95% CI based on 1,000 bootstrap replicates.

### Selection of Clinically Actionable Thresholds

We calculated sensitivities, specificities, positive predictive values, and negative predictive values across the entire spectrum of EDI thresholds. In consultation with intensivists on our research team (TSV and MWS), we identified two clinically actionable thresholds: one for identifying high-risk patients who will likely need ICU-level care (based on the EDI throughout the hospitalization) and one for identifying low-risk patients who may be appropriate for lower-acuity care (based on the 48-hour analysis).

### Software

We used R 3.6.0 for all analyses, as well as the pROC package and code adapted from the rms package.^17–19^ We have made our statistical code available at https://github.com/ml4lhs/edi_validation.

## RESULTS

We identified 392 hospitalizations for 369 patients with COVID-19 who met inclusion criteria for our study cohort. Two patients had 3 hospitalizations and 19 patients had 2 hospitalizations. At the hospitalization level (n=392), the median age of patients was 64 years (IQR 53-75), 169 (43%) were women and 168 (43%) were African Americans (**Table 1**). The composite adverse outcome occurred in 103 (26%) out of the 392 hospitalizations with a median length of follow-up of 5.3 days (IQR 2.9-10.7, max 49). The outcome occurred at a median of 2.5 days (IQR 0.65-5.0, max 24) after admission. Of all hospitalizations, 88 (22%) resulted in ICU-level care, 44 (11%) in mechanical ventilation, and 35 (8.9%) in death in the hospital. Those who experienced an adverse outcome were older, more likely to be Caucasian, and more likely to have a history of cardiac arrhythmias, chronic kidney disease, congestive heart failure, depression, diabetes, hypertension, metastatic cancer, and rheumatoid arthritis or other collagen vascular diseases (all p<0.05, see Table 1).

**Table 1.**
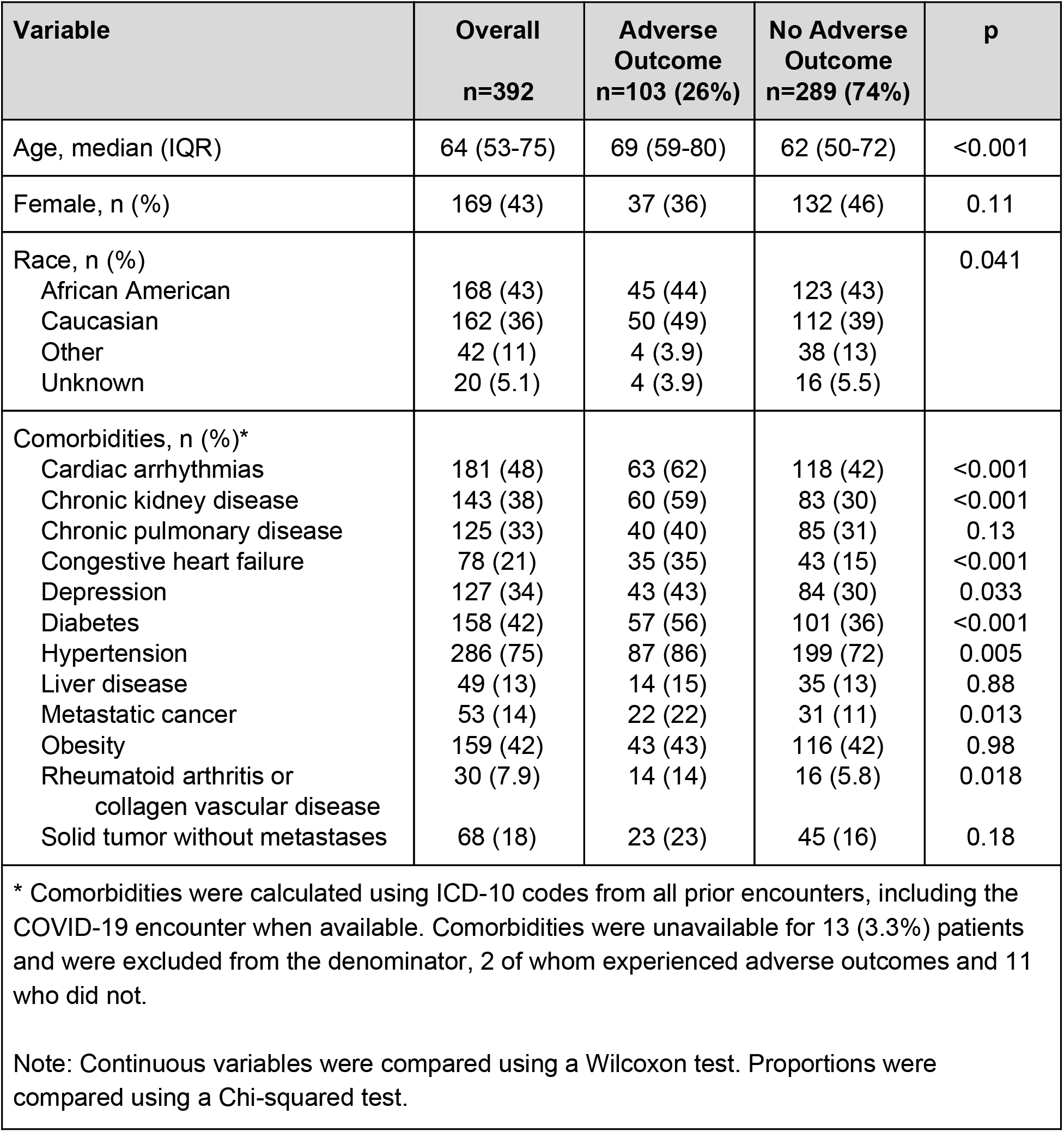
Patient characteristics, overall and stratified by adverse outcomes.

Overall, the EDI score had an AUC of 0.79 (95% CI 0.74-0.84) as a continuous predictor of risk. The performance characteristics of the EDI score are reported in **Figure 1**. Patients who met or exceeded an EDI of 68.8 had a 74% probability of experiencing the primary outcome (i.e., positive predictive value) with a sensitivity of 39%, and they comprised 14% of the study cohort. The median lead time from when the threshold was first exceeded to when the outcome occurred was 24 hours (IQR 1.4-83). The EDI slope had AUCs of 0.66 (95% CI 0.60-0.72) and 0.63 (95% CI 0.57-0.69) when assessed on a rolling basis over the prior 4 and 8 hours, respectively. **Figure 2** evaluates model calibration and demonstrates that the EDI systematically overpredicts the risk of experiencing the primary outcome if interpreted as a probability.

**Figure 1.**
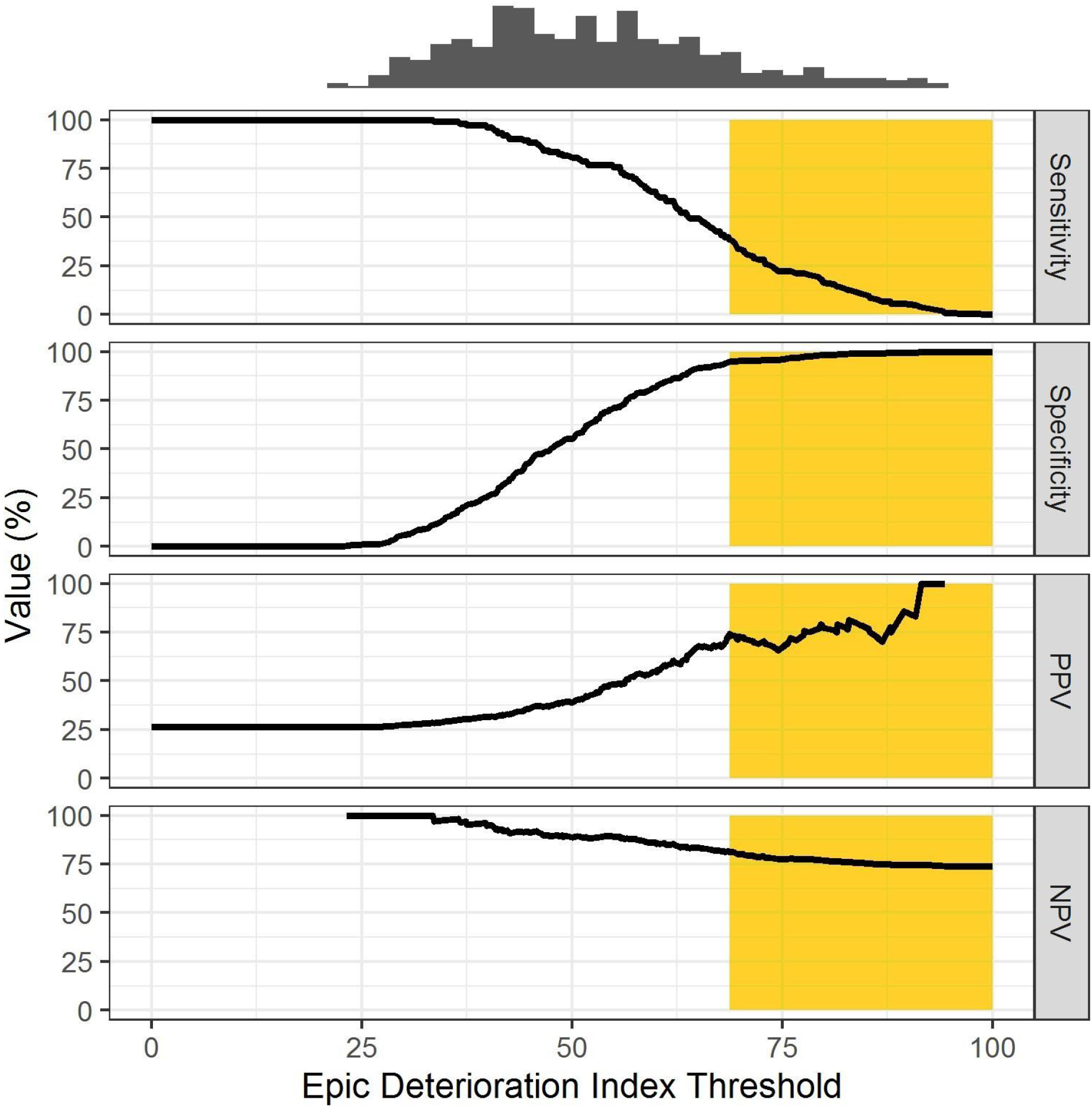
High-Risk Analysis: Plot showing the relationship between the EDI threshold and the sensitivity, specificity, positive predictive value (PPV), and negative predictive value (NPV), with a histogram demonstrating the distribution of maximum EDI per patient. High-risk area shaded in orange.

**Figure 2.**
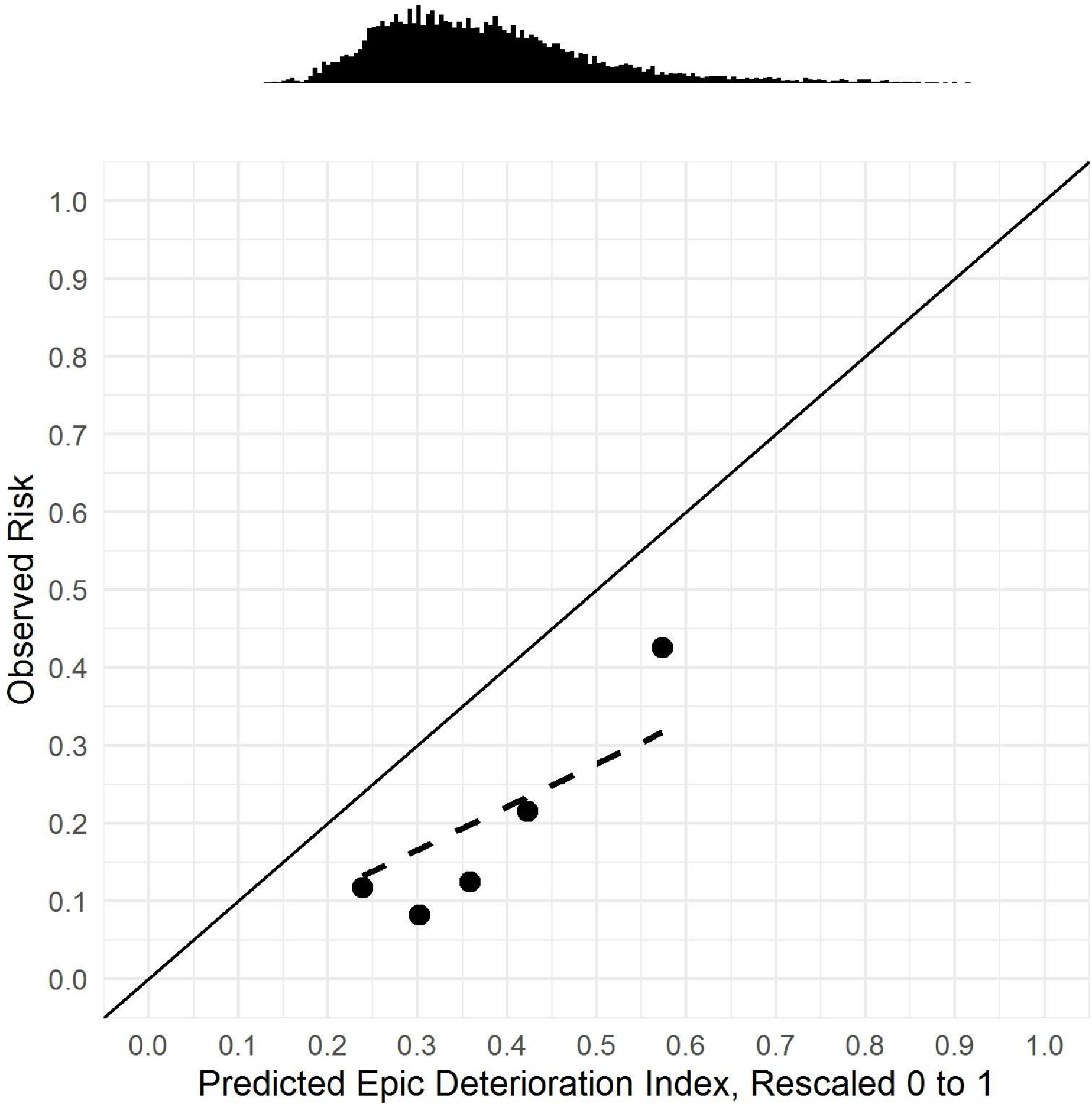
Calibration curve comparing quintiles of all predicted EDI, rescaled to 0 to 1, to the observed risk, with a line demonstrating ideal calibration (solid), a best-fit line for observed calibration (dashed), and a histogram of predicted EDI.

In an analysis of model performance by subgroup, the EDI performed similarly across all prespecified demographic subgroups (**Table 2**). In the disparate impact analyses, we found that EDI predictions did not differ by sex or race (**Appendix Table 1**). We did find higher EDI scores for patients 65 and older and those with cardiac arrhythmias, chronic kidney disease, chronic pulmonary disease, congestive heart failure, diabetes, and hypertension.

**Table 2.** Performance analysis by subgroup.

| Subgroup | Yes<br>AUC<br>(95% CI) | No<br>AUC<br>(95% CI) | p |
| --- | --- | --- | --- |
| Age ≥ 65 | 0.77 (0.69-0.84) | 0.79 (0.71-0.87) | 0.77 |
| Female | 0.83 (0.75-0.89) | 0.77 (0.70-0.84) | 0.31 |
| African American | 0.81 (0.73-0.89) | 0.78 (0.70-0.85) | 0.49 |
| Note: AUC = Area under the receiver-operating-characteristic curve. |  |  |  |

In the subset of 286 patients who had not been discharged or experienced the primary outcome at 48 hours, 55 (19%) experienced the composite outcome at some point during the remainder of their hospitalization. In this setting, the EDI had an AUC of 0.65 (95% CI 0.57-0.73). The performance characteristics of the 48-hour maximum EDI in this subset of patients are reported in **Figure 3**. A total of 14 (13%) patients who never exceeded an EDI of 37.9 in the first 48 hours of their hospitalization had a 90% probability of not experiencing the outcome (i.e., negative predictive value) for the remainder of the hospitalization (median remaining follow-up of 3.8 days [IQR 1.7-8.9]) with a sensitivity of 91% above this threshold.

**Figure 3.**
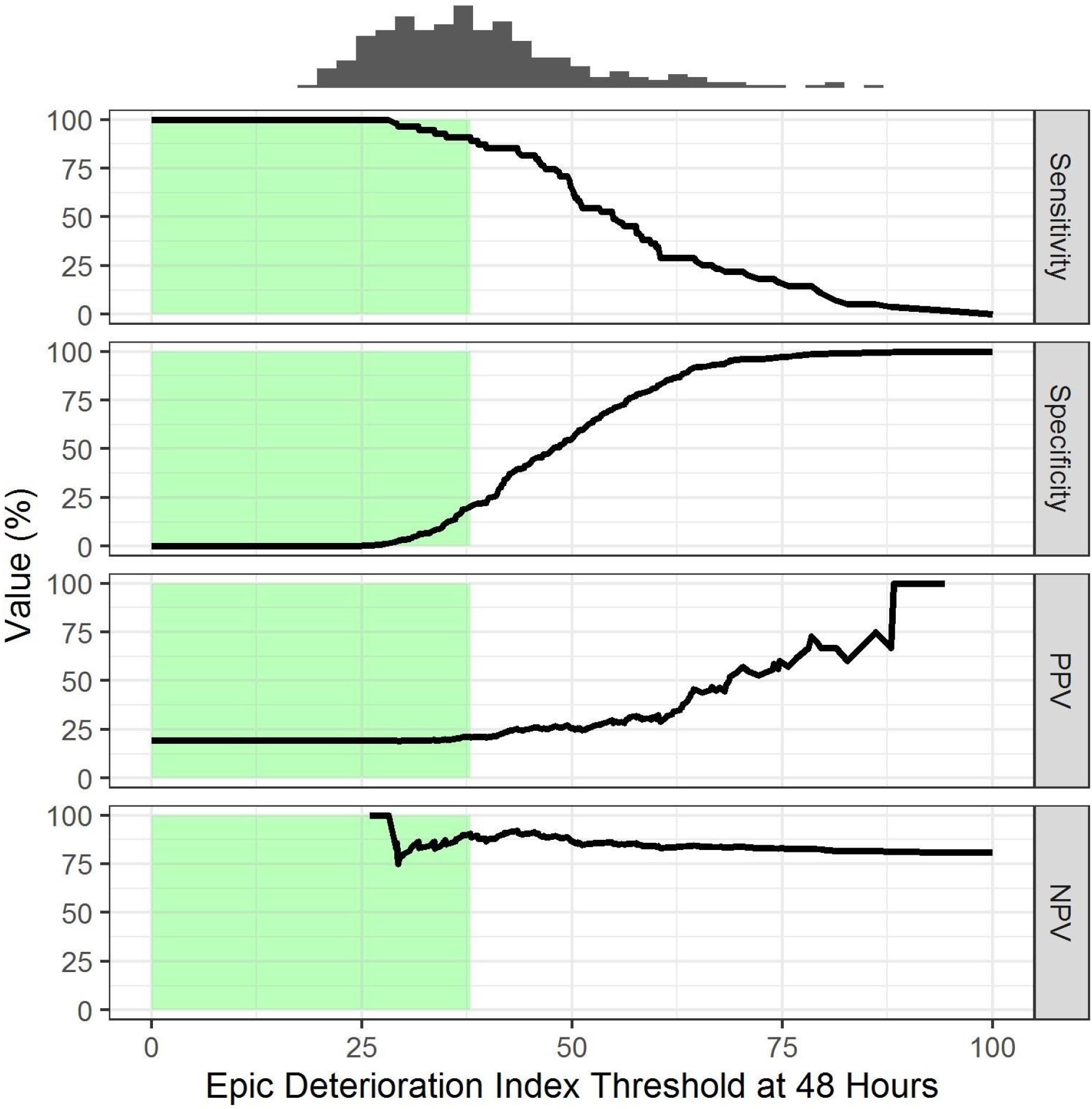
Low-Risk Analysis: Plot showing the relationship between the EDI score threshold in the first 48 hours and the sensitivity, specificity, positive predictive value (PPV), and negative predictive value (NPV), with a histogram demonstrating the distribution of maximum EDI per patient. Low-risk area shaded in green.

**Figure 4** demonstrates four examples of EDI patterns in COVID-19 patients overlaid with the identified high- and low-risk thresholds (≥ 68.8 for high-risk and <37.9 for low risk, respectively). As shown in the bottom-left and top-right panels of this figure, the EDI fluctuates substantially for individual patients with each assessment over the regular 15-minute intervals.

**Figure 4.**
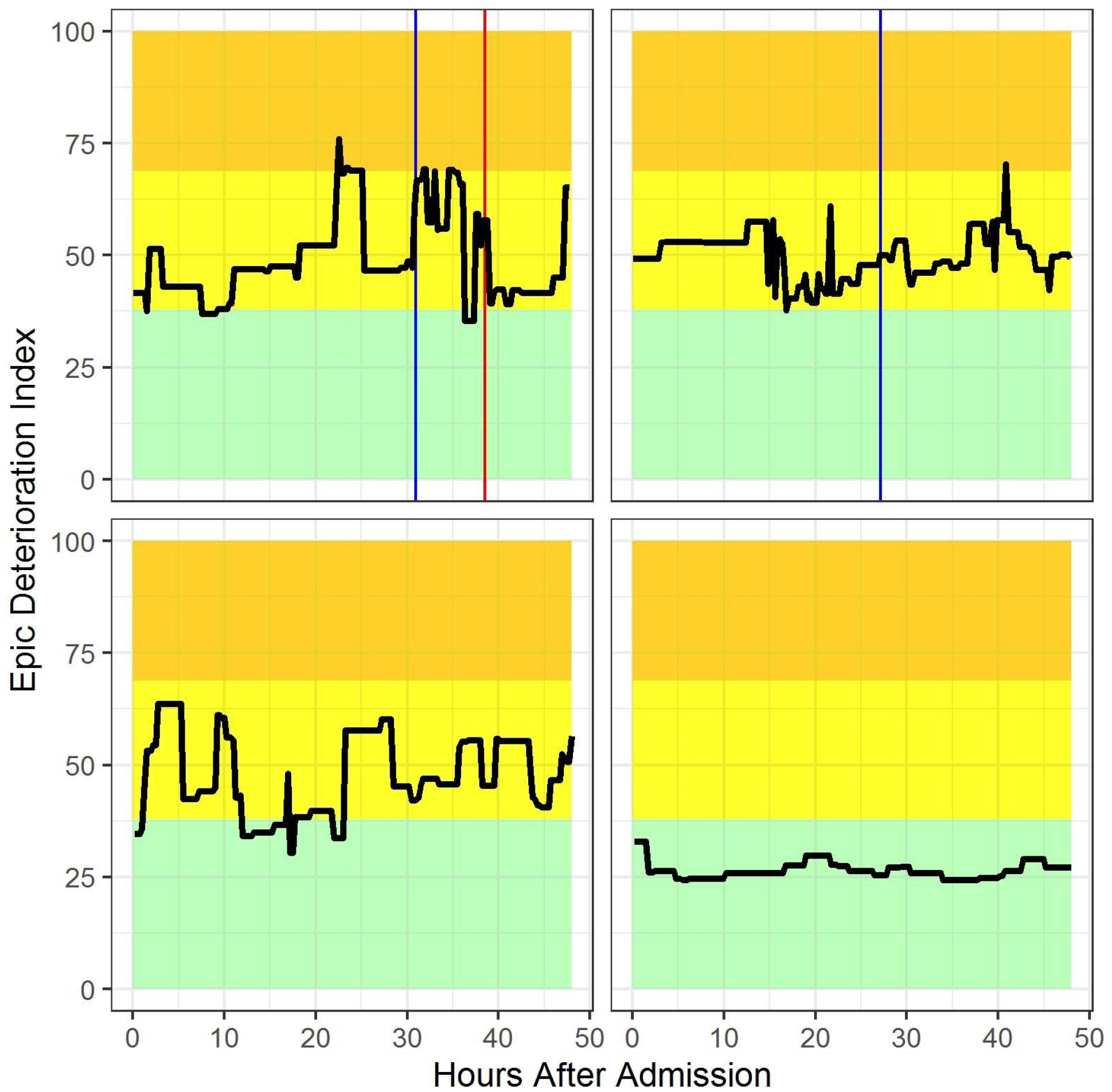
Four example patients, of whom two experienced an adverse outcome, with shaded risk thresholds. Green (EDI < 37.9): low risk, yellow: intermediate risk (≥ 37.9 to < 68.8), orange (EDI ≥ 68.8): high risk. EDI scores recorded after the primary outcome are shown in the top panels but were not used in the model validation. Blue line: transfer to intensive care unit, red line: onset of mechanical ventilation.

## DISCUSSION

Our study constitutes the first publicly reported independent validation of the EDI in any patient population. Our results suggest that the EDI exhibits fair discrimination for the prediction of adverse outcomes in a diverse COVID-19 patient population. It demonstrated good performance in identifying higher-risk patients in our cohort, identifying a small proportion of patients with a positive predictive value of 74% but a relatively low sensitivity of 39%. We found that the maximum EDI in the first 48 hours of the hospitalization may help identify a small subset of low-risk patients who may be safely transferred to lower-acuity settings, thereby conserving resources. For the vast majority of patients whose maximum EDI score falls in the intermediate-risk range, the score has limited value to guide clinical decision-making. We also noted the EDI fluctuates substantially when calculated at 15-minute intervals, in part because it only relies on the most recent value for each of the predictors. Even small changes in predictors lead to large differences in the EDI because prior normal values are ignored when a new value is obtained. Thus, we recommend the interpretation of individual EDI scores be based on whether a patient ever exceeds specific thresholds. The substantial variation in EDI scores also underscores the notion of diminishing returns when running the model so frequently. Contrary to anecdotal reports,^16^ we found the absolute EDI score to have better discrimination than the slope of the EDI score among patients with COVID-19.

The proprietary nature of the EDI raises specific ethical and clinical concerns in the setting of a pandemic, in which resources may be scarce and could be allocated to higher risk patients based on the output of this prediction model. We found no evidence that the EDI is biased against specific subgroups of vulnerable patients although the EDI was not always concordant with the observed differences in adverse outcomes (in **Table 1** as compared to **Table 2**). Although chronic pulmonary disease was not associated with an adverse outcome, the maximum EDI score was higher for patients with chronic pulmonary disease as compared to those without it. On the other hand, although adverse outcomes were more likely in Caucasians and patients with depression, metastatic cancer, and rheumatoid arthritis or other collagen vascular diseases, the maximum EDI score was not different for these subgroups. Importantly, we found no evidence for bias in the EDI score against African Americans, who are disproportionately impacted by COVID-19. The EDI score was higher in older individuals, which is not surprising because age is a component of the EDI. Even though we did not identify unfair biases in the EDI, sharing the model parameters would allow for a more complete validation and could benefit the public by enabling the model to be refined and compared with other existing predictive models for specific clinical applications such as outcomes of COVID-19.

When comparing our observed performance of the EDI in COVID-19 patients against other models reported in the literature, it is interesting to note the AUC reported in our study is much lower than other models in the COVID-19 literature. This could be due in part to overfitting of other models in the setting of relatively small sample sizes. For instance, Bai and colleagues report an AUC of 0.95 for inpatient clinical deterioration with a model developed and validated in a cohort of 133 patients with 75 predictors.^20^ By contrast, the EDI was developed on a cohort drawn from more than 130,000 hospitalizations.^6^ Our findings closely match the observed performance of other deterioration indices that have been validated in non-COVID patients. In a study of 649,418 hospitalizations, the Advanced Alert Monitor identified deteriorating patients with an AUC of 0.82.^21^ The electronic Cardiac Arrest Risk Triage (eCART) score identified deteriorating patients with an AUC of 0.80 and 0.79 in two separate evaluations.^21,22^ The Rothman index identified clinical deterioration with an AUC of 0.76.^23^

Our study should be interpreted in the context of the following limitations. Our evaluation was limited by its focus on a single academic medical center and a relatively small number of patients. However, our cohort of nearly 400 patients was diverse in sex and race and larger than many earlier reports. As compared to a recently described large cohort of 5,700 patients hospitalized with COVID-19 in New York, our study cohort had a higher proportion of African Americans (43% vs. 23%) and patients with chronic kidney disease (38% vs. 5%), congestive heart failure (21% vs. 7%), hypertension (75% vs. 57%), and similar proportions of women (43% vs. 40%), diabetes (42% vs. 34%), and obesity (42% vs. 42%).^2^ Our proposed EDI thresholds may be influenced by local factors, including patterns of COVID-19 testing, triage, and decision-making about hospital admissions and hospital-to-hospital transfers that contributed to our study cohort. These EDI thresholds should be validated in other settings to assess their generalizability.

## CONCLUSION

Despite these limitations, our findings have important implications for hospitals with access to the EDI that may be under substantial capacity constraints and strain from managing COVID-19 patients. Our study supports, in part, a role of the EDI to identify a small subset of high-risk patients who may benefit from additional resources and higher-level care and another limited subset of low-risk patients who may be cared for safely in lower-acuity settings. It also suggests opportunities to tailor and improve risk prediction for this condition beyond the EDI as data accumulate on COVID-19 patients. Finally, it indicates the need for institutions to independently validate widely used proprietary models where the vendor is commonly the only source of model validation.

## Data Availability

The data include protected health information from the University of Michigan and are not available for download. The code is available at: https://github.com/ml4lhs/edi_validation

## ACKNOWLEDGEMENTS

We would like to acknowledge Vikas Parekh, MD, Rich Medlin, MD, Max Garifullin, MSc, Justin Pestrue, MEcon, and Judy Konye, MSN for their contributions to this work.

